# Translating metagenomics into clinical practice of complex paediatric neurological presentations

**DOI:** 10.1101/2023.06.02.23290816

**Authors:** Justin Penner, Jane Hassell, Julianne R Brown, Kshitij Mankad, Nathaniel Storey, Laura Atkinson, Nisha Ranganathan, Alexander Lennon, Jack C D Lee, Dimitrios Champsas, Angelika Kopec, Divya Shah, Cristina Venturini, Garth Dixon, Surjo De, James Hatcher, Kathryn Harris, Kristian Aquilina, Maaike A. Kusters, Karyn Moshal, Delane Shingadia, Austen JJ Worth, Giovanna Lucchini, Ashirwad Merve, Thomas S Jacques, Alasdair Bamford, Marios Kaliakatsos, Judith Breuer, Sofia Morfopoulou

**Affiliations:** Great Ormond Street Hospital for Children NHS Foundation Trust, Department of Paediatric Infectious Diseases, London, UK; Great Ormond Street Hospital for Children NHS Foundation Trust, Department of Paediatric Neurology, London, UK; Great Ormond Street Hospital for Children NHS Foundation Trust, Department of Microbiology, Virology, and Infection Prevention & Control, London, UK; Great Ormond Street Hospital for Children NHS Foundation Trust, Department of Radiology, London, UK; Barts Health NHS Trust, Department of Virology East & Southeast London Pathology Partnership, London, UK; Great Ormond Street Hospital for Children NHS Foundation Trust, Department of Paediatric Neurosurgery, London, UK; Great Ormond Street Hospital for Children NHS Foundation Trust, Department of Paediatric Immunology, London, UK; Great Ormond Street Hospital for Children NHS Foundation Trust, Department of Paediatric Haematology and Bone Marrow Transplant, London, UK; Histopathology Department, Great Ormond Street Hospital for Children NHS Foundation Trust, London, UK; Developmental Biology and Cancer Department, Great Ormond Street Institute of Child Health, University College London, London, UK; UCL Great Ormond Street Institute of Child Health, London, UK; Infection, Immunity and Inflammation Department, GOS Institute of Child Health, University College London, London, UK; Section for Paediatrics, Department of Infectious Diseases, Faculty of Medicine, Imperial College London, London, UK

## Abstract

**Background:** Atypical or complex paediatric neurological presentations are common clinical conundrums and often remain undiagnosed despite extensive investigations. This is particularly pronounced in immunocompromised patients. Here we show that clinical metagenomics (CMg) is a valuable adjunct diagnostic tool to be used by neuro-infection multidisciplinary teams (MDTs).

**Methods:** We included patients referred to the Great Ormond Street Hospital neuro-infection MDT in whom diagnostic uncertainty remained, despite a standardised comprehensive set of investigations, and who were referred for untargeted CMg on brain tissue and/or cerebrospinal fluid (CSF). In a retrospective review, two clinicians independently assessed whether CMg in conjunction with the MDT resulted in a change of management.

**Findings:** 60 undiagnosed patients met the inclusion criteria. We detected the causative pathogen by CMg in 14/60 (23%), with 12/36 patients known to be immunocompromised. CMg results, even when negative, informed patient care, resulting in changes in clinical management in 42/57 (74%). Six patients had unexpected findings of pathogens not identified on prior samples. In four patients, the pathogen was found solely in the brain biopsy and was absent from all other specimens, including CSF.

**Interpretation:** CMg is particularly useful when conventional diagnostic techniques for meningoencephalitis are exhausted and proved to be an important diagnostic tool for immunocompromised patients. CMg provided increased reassurance against an infective aetiology prior to recommending immunosuppressive or immunomodulatory treatment. Specialised MDTs should advocate for early brain biopsies and routine CMg in an experienced laboratory for undiagnosed complex neurological cases affecting immunocompromised patients.

## Introduction

Paediatric meningoencephalitis is a complex diagnosis with multiple infective and non-infective aetiologies^1^. More than 50% of cases remain undiagnosed despite improvements in diagnostic techinques^2,3^. The incidence of meningoencephalitis in the US and England has increased over the past two decades, likely secondary to increased clinical use of immunomodulatory therapies and advances in transplant medicine^3^. Immunocompromised paediatric patients are at higher risk of neurological compromise, a result of difficult-to-diagnose, sometimes novel pathogens, which can cause neuroinflammation even with low infection burden^1^.

To address this challenge, we implemented routine clinical metagenomics (CMg) for children with complex neurologic presentations where the diagnosis of central nervous system (CNS) infection, neuro-inflammation, or neurotoxicity was uncertain. The use of CMg (http://www.labs.gosh.nhs.uk/laboratory-services/microbiology-virology-and-infection-control/metagenomics-pathogen-detection) was established in 2014 as a collaboration between Great Ormond Street Hospital (GOSH) and University College London (UCL) but has since evolved into a clinical service aiming for UKAS accreditation.

In parallel with the laboratory metagenomics service (**Figure 1**), a regular neuro-infection multidisciplinary team (MDT) (**Figure 2**) was established to discuss the investigation, management, and outcomes of children with complex or undiagnosed paediatric CNS pathology. Membership of the MDT includes physicians, clinical nurse specialists, and clinical scientists from infectious disease, neurology, laboratory and clinical immunology, microbiology/virology, neuroradiology, rheumatology, and neurosurgery.

**Figure 1.**
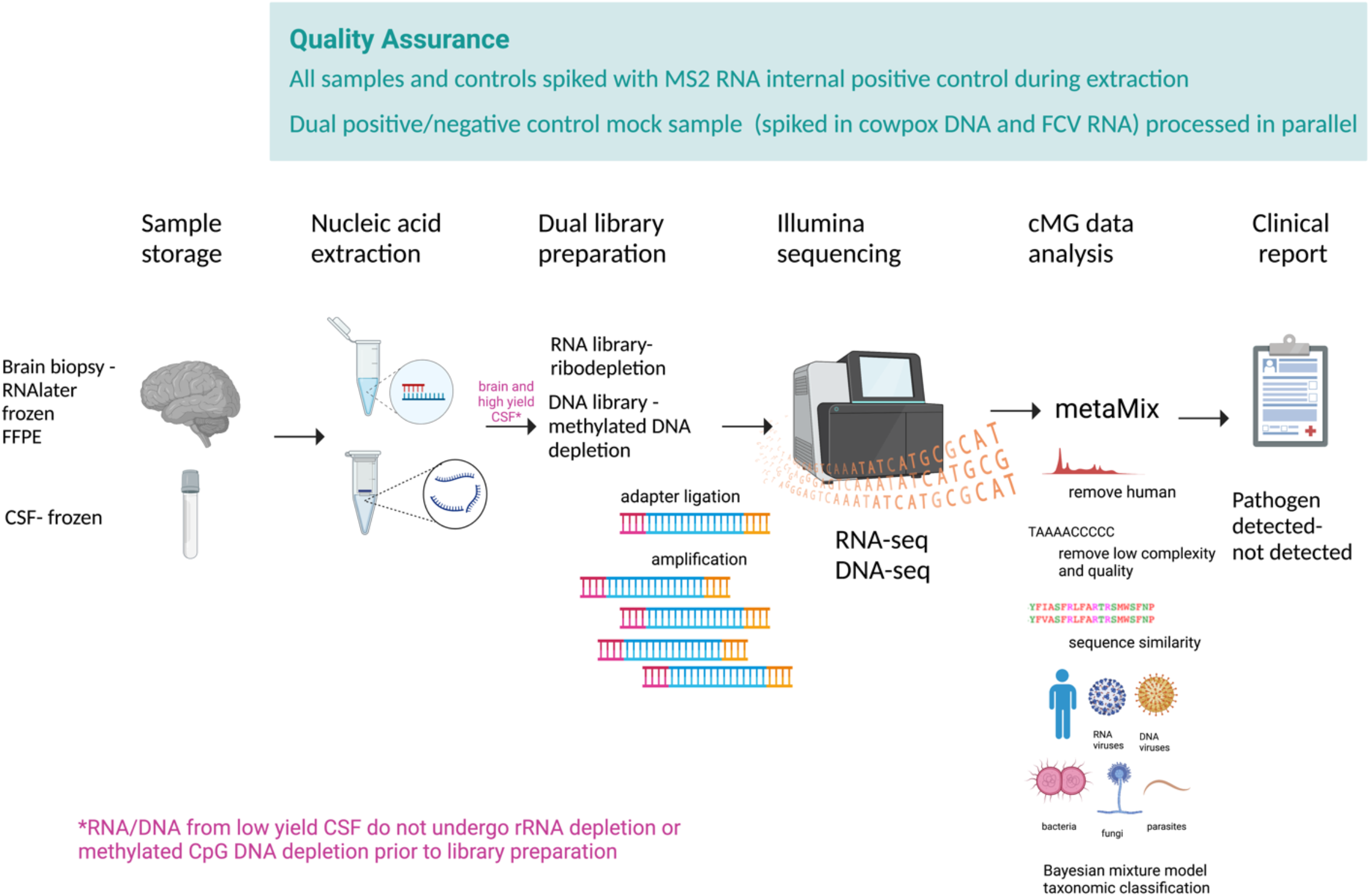
Clinical Metagenomics Pipeline. CSF: cerebrospinal fluid; CMg: clinical metagenomics; FFPE: formalin-fixed paraffin-embedded tissue. Figure created with BioRender.

**Figure 2.**
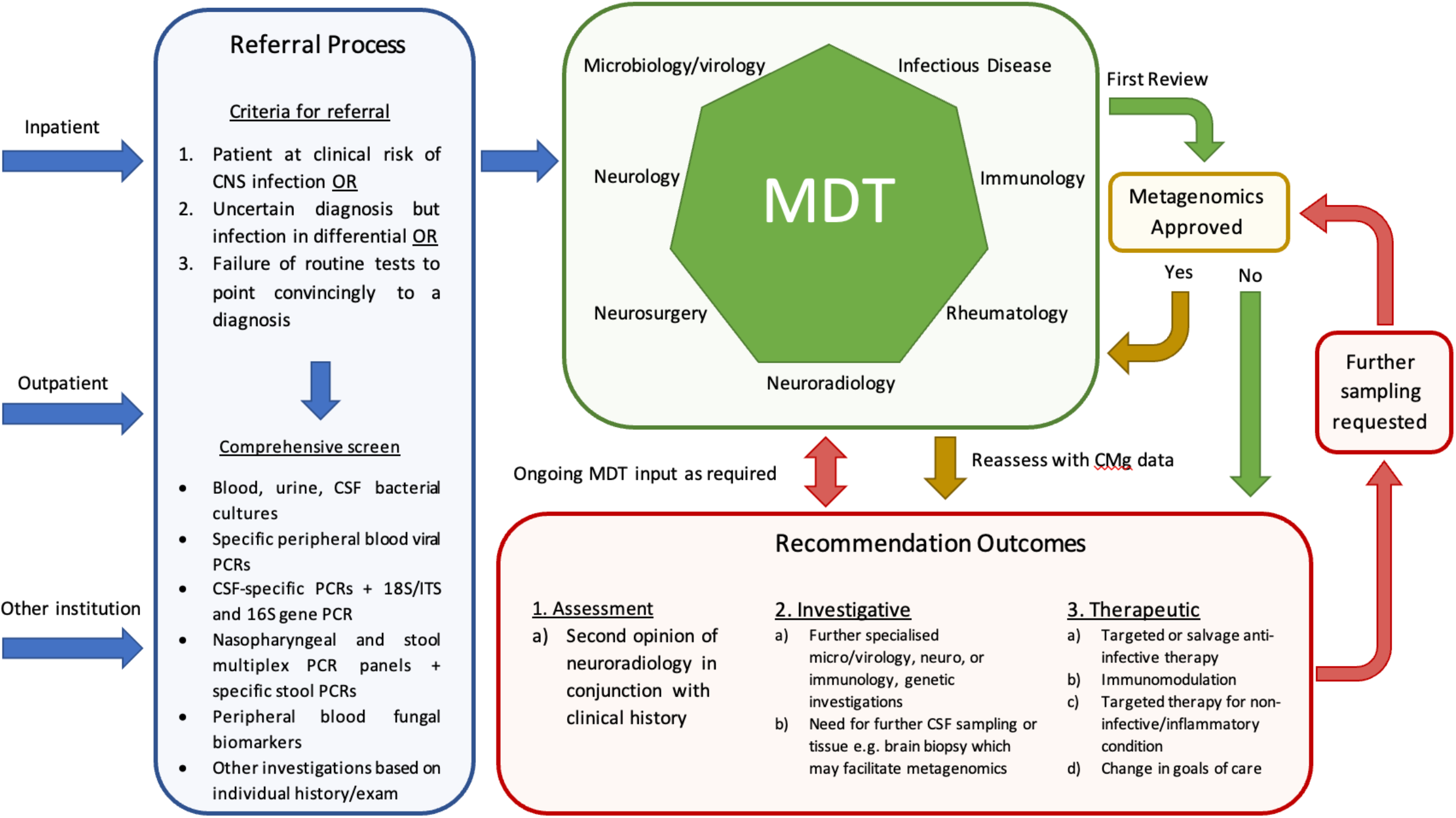
Great Ormond Street Neuro-infection MDT clinical pathway CMg: clinical metagenomics; CNS: central nervous system; CSF: cerebrospinal fluid; ITS: internal transcribed spacer; MDT: multidisciplinary team; PCR: polymerase chain reaction

Here, we show the impact a CMg service integrated with a neuro-infection MDT has made on a case-series of 60 paediatric patients at a tertiary and quaternary paediatric hospital in London, United Kingdom.

## Methods

### Ethics

The neuro-infection MDT service evaluation was registered as a clinical audit with the GOSH NHS Trust. Upon data extraction, patient information was deidentified and rare diseases classified into broad diagnostic categories to preserve anonymity.

### Inclusion in the current study

For the current study, we discuss only the GOSH patients referred through the neuro-infection MDT. CMg referral was decided by the MDT, based on the following three criteria: 1) Patient at clinical risk of CNS infection 2) Uncertain neurological diagnosis, but infection in the differential diagnosis 3) Failure of routine tests to point convincingly to a diagnosis. Retrospective data on patients that had died or were discussed in the MDT post-mortem were included as applicable.

### Data Extraction

Data were extracted from electronic medical records (EPIC®). MDT outcomes were assessed and categorised independently by two reviewers (JP & JHas). A change in clinical management was determined based on the MDT input, considering the impact of both positive and negative CMg results. When there was disagreement between reviewers, a third reviewer independently examined patient data to mediate a decision. Inborn errors of immunity were classified based on the International Union of Immunological Studies 2022 classification^4^.

### Metagenomics

Patients referred for CMg between 2014 and 2018 had ribonucleic acid-sequencing (RNA-seq) on brain biopsies (Cases 1-5, 18-23, 25-33, 46). Patients from 2019 onwards had both deoxyribonucleic acid-sequencing (DNA-seq) and RNA-seq on their brain biopsy and/or CSF, as the service expanded from viral detection in brain biopsy to universal pathogen detection in brain biopsy or cerebrospinal fluid (CSF).

As part of the GOSH neuro-infection MDT service, all patients undergo a standard comprehensive set of targeted microbiology and neurology investigations prior to CMg (**Supplementary Table 1**). All remaining infection investigations are recommended based on the individual history and clinical findings on examination of the patient. Confirmation of CMg findings with specific polymerase chain reaction (PCRs) on available tissue and/or body fluids and other tests, such as immunohistochemistry, is undertaken as required.

CSF is stored at -80°C within 24 hours of collection to minimise RNA degradation. Fresh brain tissue biopsies are placed directly into the preservative RNALater at the point of collection. In situations when RNALater is not available, fresh brain tissue biopsy is stored immediately at -80°C and upon receipt into the laboratory is placed directly into RNAlater ICE for 24 hours at -20 °C. Some retrospectively referred cases are analysed from formalin fixed paraffin embedded (FFPE) brain tissue.

Untargeted Illumina metagenomics (DNA-seq) and metatranscriptomics (RNA-seq) are undertaken in line with recommended guidelines for clinical metagenomics^5,6^. Details of current nucleic acid extraction and sequencing library preparation protocols can be found in the **Supplementary Methods**.

The limit of detection was established using model organisms. For RNA organisms the limit of detection is equivalent to targeted real-time PCR in both tissue and CSF (**Supplementary Methods**). For DNA organisms the limit of detection in CSF is equivalent to targeted real-time PCR. In tissue the limit of detection of DNA organisms is approximately 100-fold less sensitive than targeted real-time PCR but comparable to 16S rRNA gene pan-bacterial PCR used clinically for diagnosis of culture-negative infections^7^ (**Supplementary Methods**).

### Computational analysis

The 2014-2018 pipeline has been reported previously^8–11^. Since 2019, pre-processing consists of filtering steps and a similarity search (**Supplementary Methods** for details). DNA and RNA sequence data are taxonomically classified with metaMix^8^ (version 0.4). metaMix-protein is optimal for RNA virus detection and metaMix-nucl is best suited for DNA microbes speciation. The latter is used for DNA-seq while both modes are used for RNA-seq. The MS2 phage RNA internal control is expected to be detected in every sample and control and feline calicivirus RNA and cowpox DNA should be detected in the controls. The metaMix statistical output is examined for false positives, including assessment of the posterior probability of species presence, the read classifications probabilities and the Bayes Factor of species being present versus being absent. The genome coverage and depth for species of interest are visualised by aligning all non-human reads to the relevant reference genome. Low level kit or skin flora contaminants that are usually present are flagged using the controls for reference. Since 2019, the average DNA sequencing depth for brain biopsies was 57 million 81-bp pairs and 23 million pairs for CSF, while RNA-seq depth was 51 million and 28 million pairs for brain and CSF respectively (**Supplementary Table 2**). For the typical dataset, over 98% of the sequences generated are human, and thus bioinformatically filtered and not examined in any downstream analyses.

## Results

### Demographics

From 2014 to 2022, the GOSH metagenomics pathogen detection service processed 195 samples from 178 patients. To determine the impact of CMg on patient management, we focused on patients with recorded demographic and clinical data, and therefore all externally referred non-GOSH patients (108/178) were excluded from the analyses.70 (40%) were GOSH patients, of whom 10 were referred to the service for a non-neurological condition. 60 patients were evaluated in the neuro-infection MDT and referred for CMg.

Of the 60 patients referred by the MDT, 52% (n=31) were male with an average age of 7 years [range 0.25-17 years] (**Table 1**). 23% (n=14) were previously healthy. 60% (n=36) were immunocompromised at baseline; 39% (n=14) of immunocompromised patients were post-haematopoietic stem cell transplant (HSCT) and 6% (n=2) post CAR-T therapy (**Table 1**). One patient was post-thymus transplant. 56% (n=20) of the immunocompromised patients had a diagnosed inborn error of immunity, 31% (n=11) had a primary oncologic diagnosis, 11% (n=4) had rheumatologic/inflammatory conditions, and 3% (n=1) had a primary metabolic condition. One patient had received a HSCT more than ten years previously, with immune-reconstitution and was off immunosuppressive medications, and another patient had isolated alopecia at the time of presentation; both were deemed immunocompetent. Of the 24 immunocompetent patients, six had prior neurological conditions, not all with defined aetiologies. None of these six patients were on immunosuppressive medications at the time of presentation or known to be immunocompromised in association with their underlying neurological condition. One patient with spinal muscular atrophy had recently received gene therapy (antisense oligonucleotide inhibitor). One patient gave a prior history of an infective encephalitis, in this case herpes simplex virus type 1 (HSV-1). Detailed demographic details, presenting neurological and neuroradiological features are shown in **Table 2**.

**Table 1.** (a) Patients’ demographic and clinical characteristics. (b) Sampling type and CMg findings.

| Demographic and Clinical Characteristics | N | Percentage % |
| --- | --- | --- |
| **Patients** | 60 |  |
| **Age** |  |  |
| Median/Range | 7 / 0.25-17 y |  |
| 0-1 | 3 | 5% |
| 1-5 | 24 | 40% |
| 6-10 | 15 | 25% |
| 11-15 | 16 | 27% |
| >=16 | 2 | 3% |
| **Sex** |  |  |
| Female | 29 | 48% |
| Male | 31 | 52% |
| **Outcome** |  |  |
| Hospitalized | 60 | 100% |
| Died | 15 | 25% |
| **Health at baseline** |  |  |
| Immunocompromised/suppressed | 36 | 60% |
| Primary Immune deficiency | 16 | 35% |
| Malignancy | 9 | 20% |
| Post HSCT/CART/thymus transplant | 11 | 24% |
| Known rheumatological/inflammatory conditions | 3 | 7% |
| Previously Healthy | 14 | 23% |

b.
| Sampling type and CMg report | N | Percentage % |
| --- | --- | --- |
| **Patients** | 60 |  |
| **CNS Sampling** |  |  |
| CSF | 21 | 35% |
| CNS Tissue | 35 | 58% |
| CSF and CNS Tissue | 4 | 7% |
| **CMg report** |  |  |
| Positive - Pathogenic | 14 | 23% |
| Positive – non pathogenic | 4 | 7% |
| Negative | 42 | 70% |

CART: chimeric antigen T-cell receptor; CMg: clinical metagenomics; CNS: central nervous system; CSF: cerebrospinal fluid; HSCT: haematopoietic stem cell transplant;

**Table 2:**
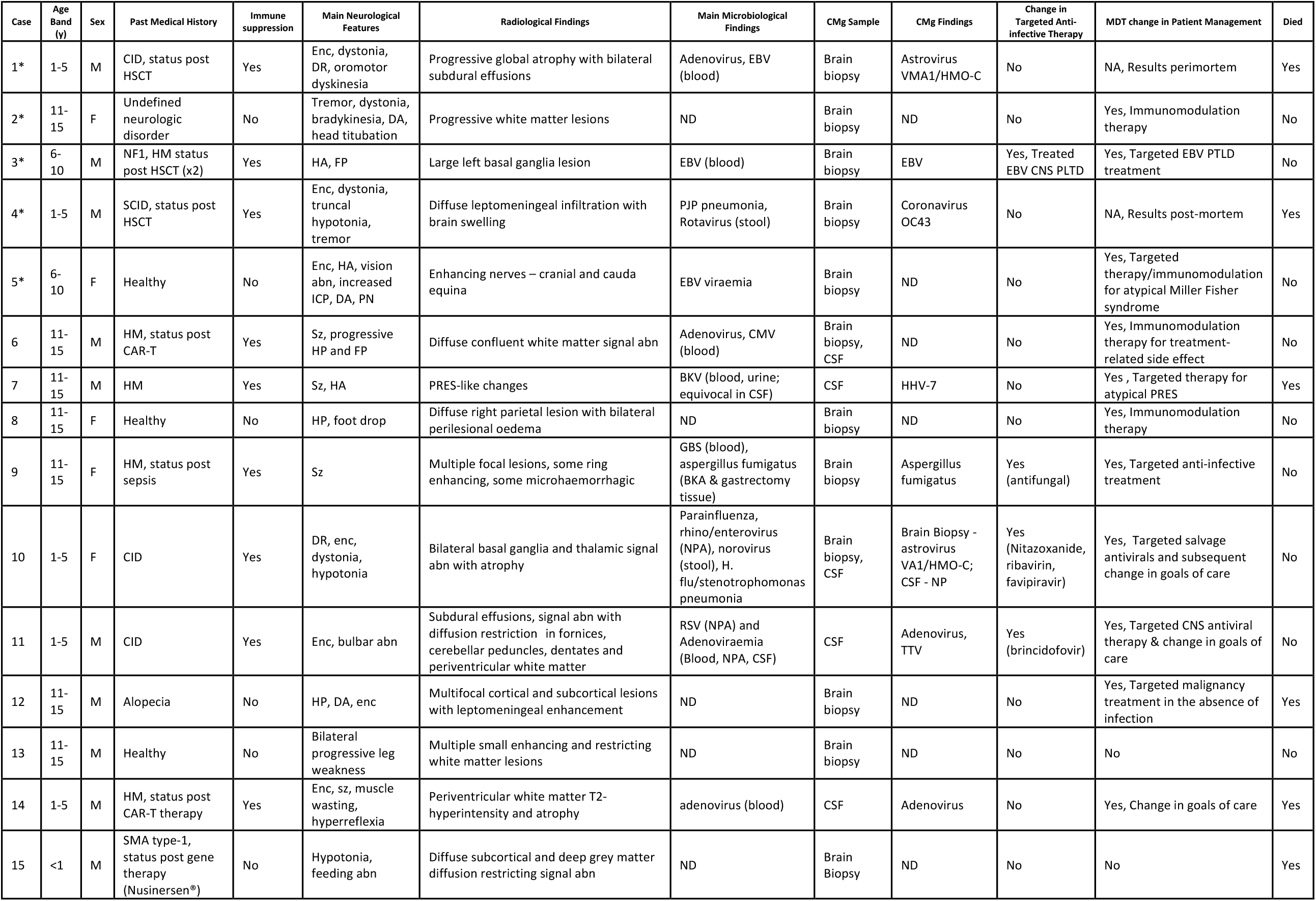
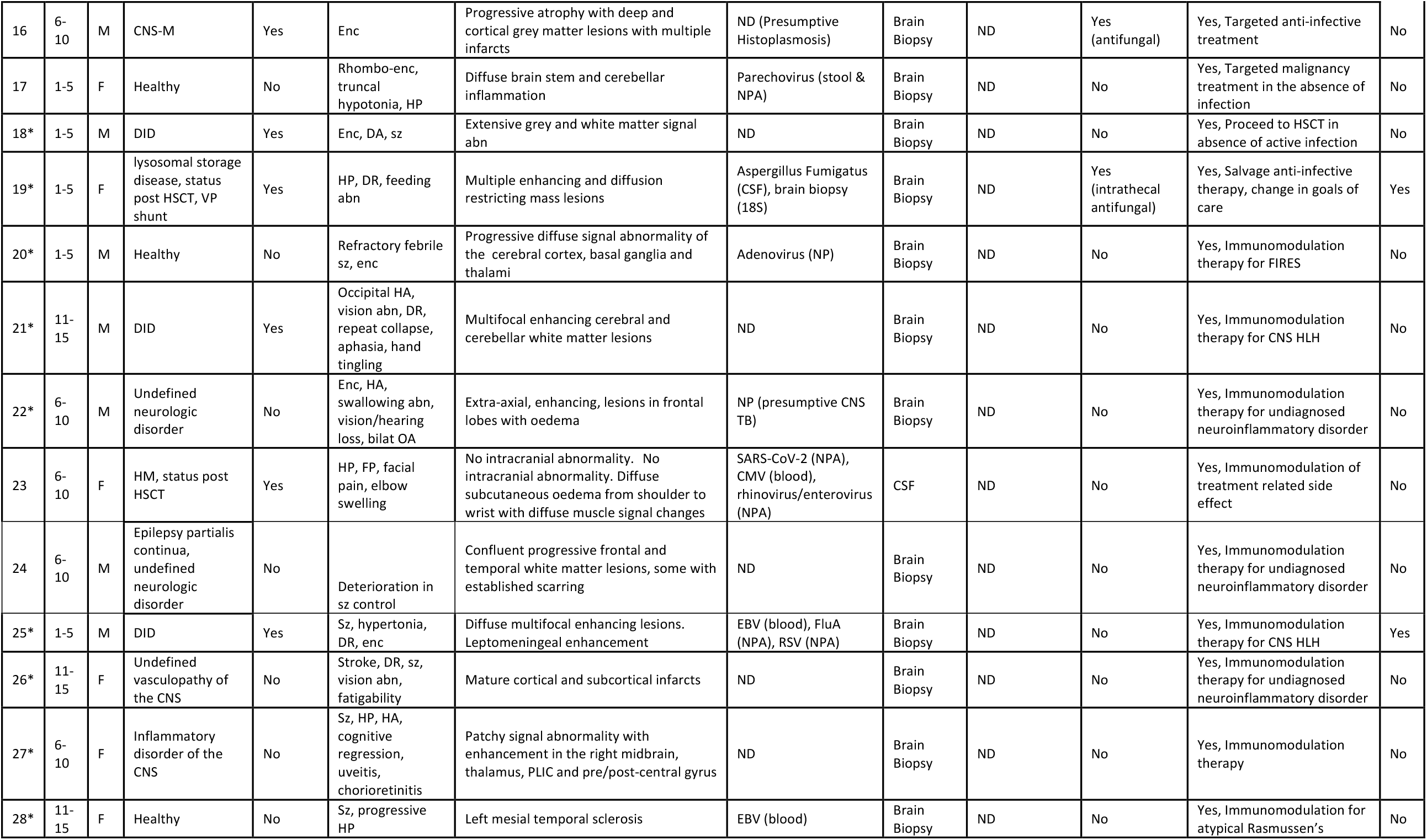
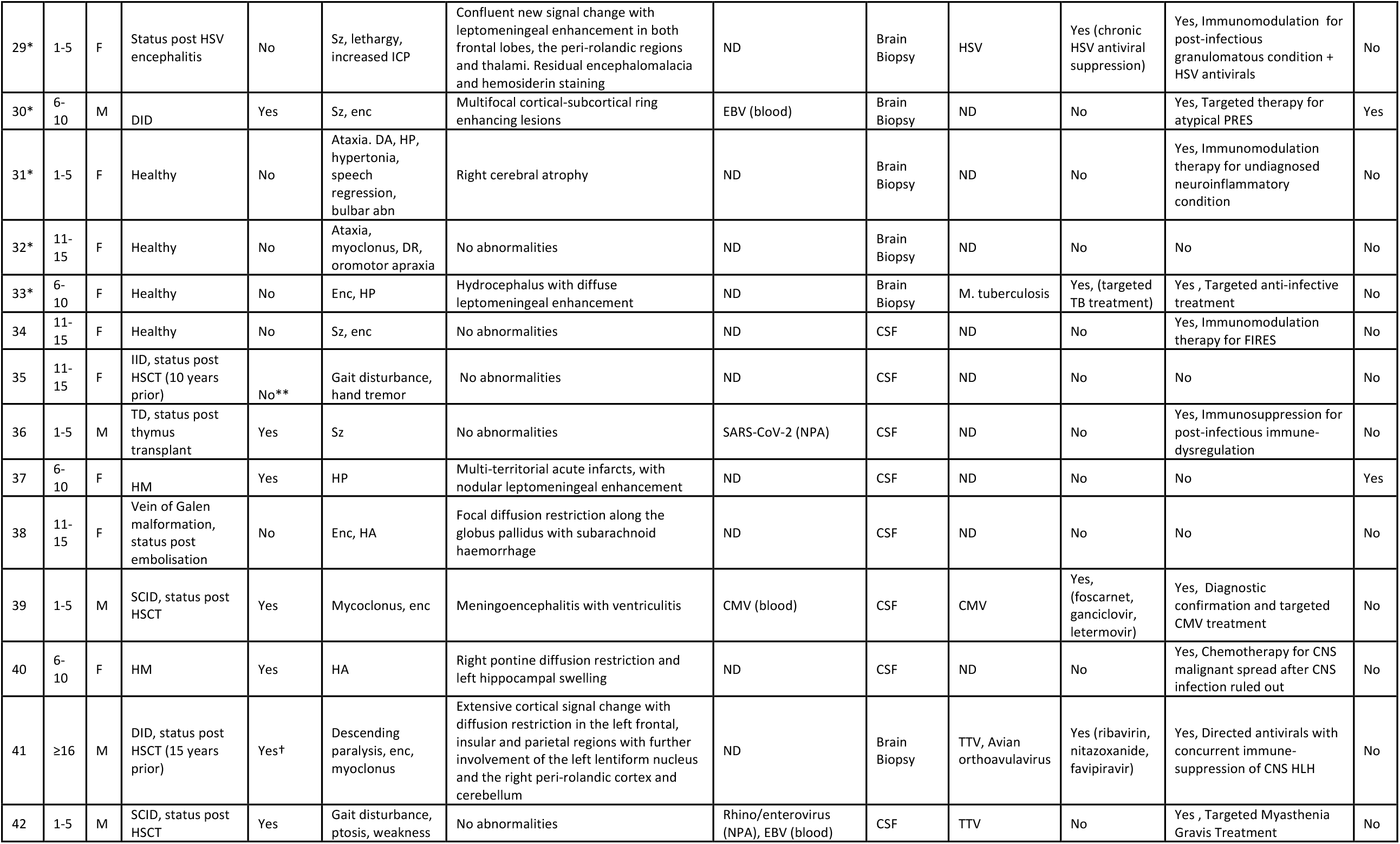
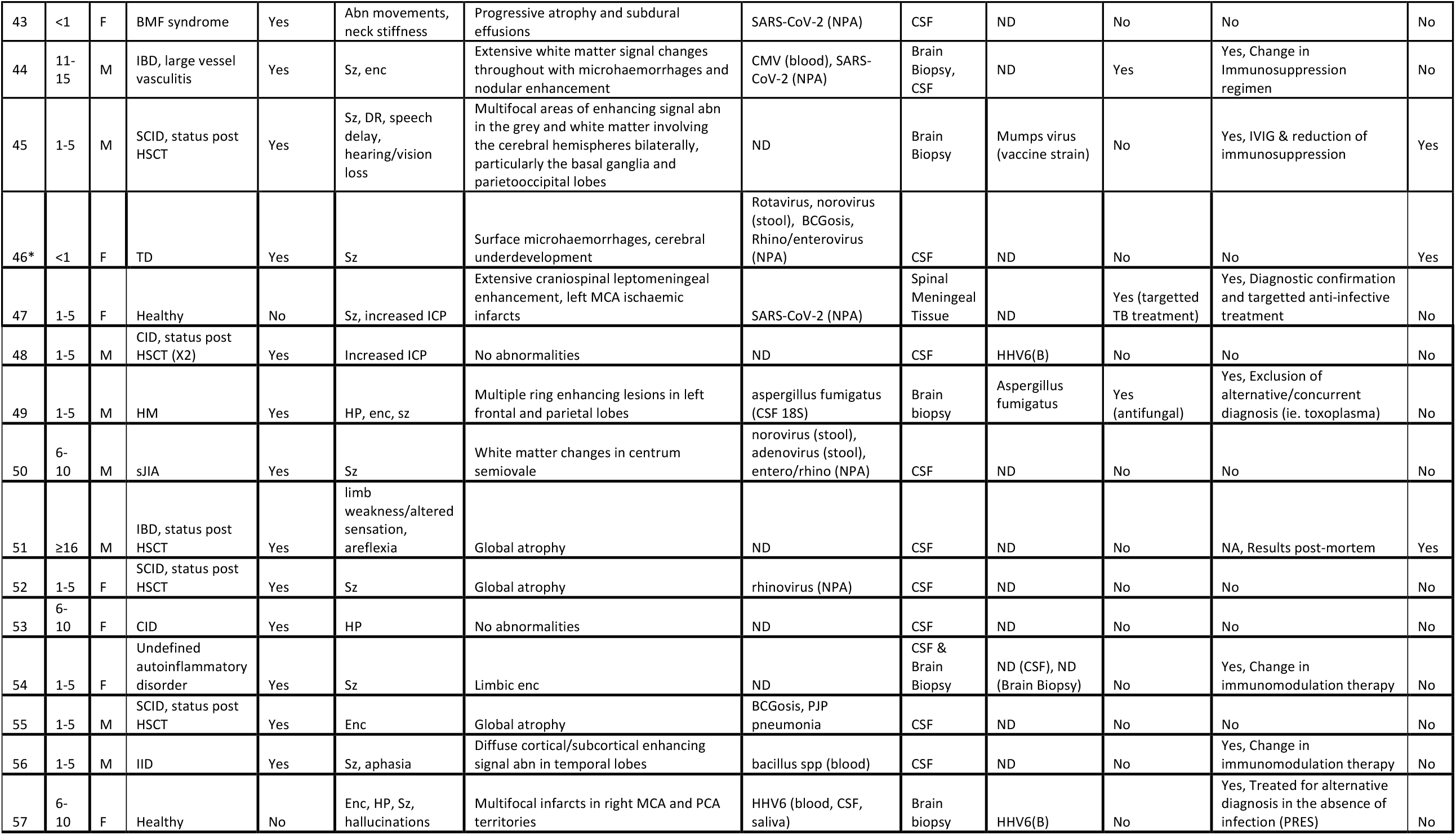
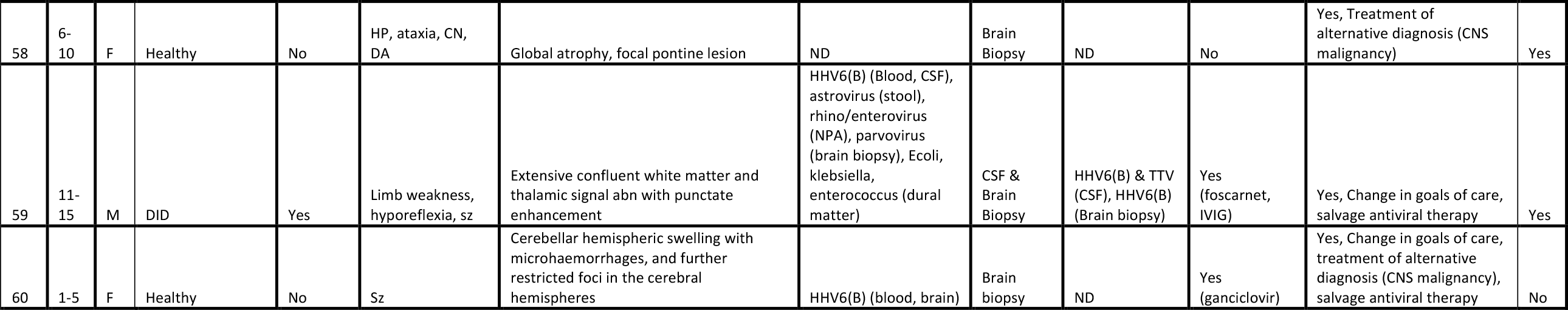
Demographic and clinical features (with radiological, microbiological and metagenomics results) of patients referred for metagenomics with related clinical management outcomes of patients managed in neuro-infection multidisciplinary team. *pre-2019 CMg pipeline **fully engrafted and off immunosuppression †poor immune reconstitution Abn: abnormalities; BKA: below knee amputation; BMF: bone marrow failure; CAR-T: chimeric antigen receptor t-cell therapy; CID: combined immunodeficiency; CMg: clinical metagenomics; CN: cranial neuropathy; CNS-M: central nervous system malignancy; DA: dysarthria; DID: disorder of immune dysregulation; DR: developmental/behavioural regression; Enc: encephalopathic; FIRES: febrile infection-related epilepsy syndrome; FP: facial palsy; GBS: group B streptococcus; HA: headache; HLH: haemophagocytic lymphohistiocytosis; HM: haematologic malignancy; HP: hemiparesis/hemiparalysis; HSCT: haematopoietic stem cell transplant; IBD: inflammatory bowel disease; ICP: intracranial pressure; IID: innate immune deficiency; MCA: middle cerebral artery; MDRTB: multi-drug resistant tuberculosis; MDT: multidisciplinary team; ND: nil detected; NF1: neurofibromatosis type-1; NPA: nasopharygeal aspirate; OA: optic atrophy; PCA: posterior cerebral artery; PJP: pneumocystis jirovecii pneumonia; PLIC: posterior limb of internal capsule; PN: peripheral neuropathy; PRES: posterior reversible encephalopathy syndrome; PTLD: post-transplant lymphoproliferative disorder; SCID: severe combined immunodeficiency; sJIA: systemic onsert juvenile idiopathic arthritis; SMA: spinal muscular atrophy; Sz: seizures; TB: tuberculosis; TD: thymic disorder; VP; ventriculoperitoneal

| Case | Age Band (y) | Sex | Past Medical History | Immune suppression | Main Neurological Features | Radiological Findings | Main Microbiological Findings | CMg Sample | CMg Findings | Change in Targeted Anti-infective Therapy | MDT change in Patient Management | Died |
| --- | --- | --- | --- | --- | --- | --- | --- | --- | --- | --- | --- | --- |
| 1* | 1-5 | M | CID, status post HSCT | Yes | Enc, dystonia, DR, oromotor dyskinesia | Progressive global atrophy with bilateral subdural effusions | Adenovirus, EBV (blood) | Brain biopsy | Astrovirus VMA1/HMO-C | No | NA, Results perimortem | Yes |
| 2* | 11-15 | F | Undefined neurologic disorder | No | Tremor, dystonia, bradykinesia, DA, head titubation | Progressive white matter lesions | ND | Brain biopsy | ND | No | Yes, Immunomodulation therapy | No |
| 3* | 6-10 | M | NF1, HM status post HSCT (x2) | Yes | HA, FP | Large left basal ganglia lesion | EBV (blood) | Brain biopsy | EBV | Yes, Treated EBV CNS PLTD | Yes, Targeted EBV PTLD treatment | No |
| 4* | 1-5 | M | SCID, status post HSCT | Yes | Enc, dystonia, truncal hypotonia, tremor | Diffuse leptomeningeal infiltration with brain swelling | PJP pneumonia, Rotavirus (stool) | Brain biopsy | Coronavirus OC43 | No | NA, Results post-mortem | Yes |
| 5* | 6-10 | F | Healthy | No | Enc, HA, vision abn, increased ICP, DA, PN | Enhancing nerves – cranial and cauda equina | EBV viraemia | Brain biopsy | ND | No | Yes, Targeted therapy/immunomodulation for atypical Miller Fisher syndrome | No |
| 6 | 11-15 | M | HM, status post CAR-T | Yes | Sz, progressive HP and FP | Diffuse confluent white matter signal abn | Adenovirus, CMV (blood) | Brain biopsy, CSF | ND | No | Yes, Immunomodulation therapy for treatment-related side effect | No |
| 7 | 11-15 | M | HM | Yes | Sz, HA | PRES-like changes | BKV (blood, urine; equivocal in CSF) | CSF | HHV-7 | No | Yes, Targeted therapy for atypical PRES | Yes |
| 8 | 11-15 | F | Healthy | No | HP, foot drop | Diffuse right parietal lesion with bilateral perilesional oedema | ND | Brain biopsy | ND | No | Yes, Immunomodulation therapy | No |
| 9 | 11-15 | F | HM, status post sepsis | Yes | Sz | Multiple focal lesions, some ring enhancing, some microhaemorrhagic | GBS (blood), aspergillus fumigatus (BKA & gastrectomy tissue) | Brain biopsy | Aspergillus fumigatus | Yes (antifungal) | Yes, Targeted anti-infective treatment | No |
| 10 | 1-5 | F | CID | Yes | DR, enc, dystonia, hypotonia | Bilateral basal ganglia and thalamic signal abn with atrophy | Parainfluenza, rhino/enterovirus (NPA), norovirus (stool), H. flu/stenotrophomonas pneumonia | Brain biopsy, CSF | Brain Biopsy - astrovirus VA1/HMO-C; CSF - NP | Yes (Nitazoxanide, ribavirin, favipiravir) | Yes, Targeted salvage antivirals and subsequent change in goals of care | No |
| 11 | 1-5 | M | CID | Yes | Enc, bulbar abn | Subdural effusions, signal abn with diffusion restriction in fornices, cerebellar peduncles, dentates and periventricular white matter | RSV (NPA) and Adenoviraemia (Blood, NPA, CSF) | CSF | Adenovirus, TTV | Yes (brincidofovir) | Yes, Targeted CNS antiviral therapy & change in goals of care | No |
| 12 | 11-15 | M | Alopecia | No | HP, DA, enc | Multifocal cortical and subcortical lesions with leptomeningeal enhancement | ND | Brain biopsy | ND | No | Yes, Targeted malignancy treatment in the absence of infection | Yes |
| 13 | 11-15 | M | Healthy | No | Bilateral progressive leg weakness | Multiple small enhancing and restricting white matter lesions | ND | Brain biopsy | ND | No | No | No |
| 14 | 1-5 | M | HM, status post CAR-T therapy | Yes | Enc, sz, muscle wasting, hyperreflexia | Periventricular white matter T2-hyperintensity and atrophy | adenovirus (blood) | CSF | Adenovirus | No | Yes, Change in goals of care | Yes |
| 15 | <1 | M | SMA type-1, status post gene therapy (Nusinersen®) | No | Hypotonia, feeding abn | Diffuse subcortical and deep grey matter diffusion restricting signal abn | ND | Brain Biopsy | ND | No | No | Yes |
| 16 | 6-10 | M | CNS-M | Yes | Enc | Progressive atrophy with deep and cortical grey matter lesions with multiple infarcts | ND (Presumptive Histoplasmosis) | Brain Biopsy | ND | Yes (antifungal) | Yes, Targeted anti-infective treatment | No |
| 17 | 1-5 | F | Healthy | No | Rhomb-enc, truncal hypotonia, HP | Diffuse brain stem and cerebellar inflammation | Parechovirus (stool & NPA) | Brain Biopsy | ND | No | Yes, Targeted malignancy treatment in the absence of infection | No |
| 18* | 1-5 | M | DID | Yes | Enc, DA, sz | Extensive grey and white matter signal abn | ND | Brain Biopsy | ND | No | Yes, Proceed to HSCT in absence of active infection | No |
| 19* | 1-5 | F | lysosomal storage disease, status post HSCT, VP shunt | Yes | HP, DR, feeding abn | Multiple enhancing and diffusion restricting mass lesions | Aspergillus Fumigatus (CSF), brain biopsy (18S) | Brain Biopsy | ND | Yes (intrathecal antifungal) | Yes, Salvage anti-infective therapy, change in goals of care | Yes |
| 20* | 1-5 | M | Healthy | No | Refractory febrile sz, enc | Progressive diffuse signal abnormality of the cerebral cortex, basal ganglia and thalami | Adenovirus (NP) | Brain Biopsy | ND | No | Yes, Immunomodulation therapy for FIRES | No |
| 21* | 11-15 | M | DID | Yes | Occipital HA, vision abn, DR, repeat collapse, aphasia, hand tingling | Multifocal enhancing cerebral and cerebellar white matter lesions | ND | Brain Biopsy | ND | No | Yes, Immunomodulation therapy for CNS HLH | No |
| 22* | 6-10 | M | Undefined neurologic disorder | No | Enc, HA, swallowing abn, vision/hearing loss, bilat OA | Extra-axial, enhancing, lesions in frontal lobes with oedema | NP (presumptive CNS TB) | Brain Biopsy | ND | No | Yes, Immunomodulation therapy for undiagnosed neuroinflammatory disorder | No |
| 23 | 6-10 | F | HM, status post HSCT | Yes | HP, FP, facial pain, elbow swelling | No intracranial abnormality. No intracranial abnormality. Diffuse subcutaneous oedema from shoulder to wrist with diffuse muscle signal changes | SARS-CoV-2 (NPA), CMV (blood), rhinovirus/enterovirus (NPA) | CSF | ND | No | Yes, Immunomodulation of treatment related side effect | No |
| 24 | 6-10 | M | Epilepsy partialis continua, undefined neurologic disorder | No | Deterioration in sz control | Confluent progressive frontal and temporal white matter lesions, some with established scarring | ND | Brain Biopsy | ND | No | Yes, Immunomodulation therapy for undiagnosed neuroinflammatory disorder | No |
| 25* | 1-5 | M | DID | Yes | Sz, hypertonia, DR, enc | Diffuse multifocal enhancing lesions. Leptomeningeal enhancement | EBV (blood), FluA (NPA), RSV (NPA) | Brain Biopsy | ND | No | Yes, Immunomodulation therapy for CNS HLH | Yes |
| 26* | 11-15 | F | Undefined vasculopathy of the CNS | No | Stroke, DR, sz, vision abn, fatigability | Mature cortical and subcortical infarcts | ND | Brain Biopsy | ND | No | Yes, Immunomodulation therapy for undiagnosed neuroinflammatory disorder | No |
| 27* | 6-10 | F | Inflammatory disorder of the CNS | No | Sz, HP, HA, cognitive regression, uveitis, chorioretinitis | Patchy signal abnormality with enhancement in the right midbrain, thalamus, PLIC and pre/post-central gyrus | ND | Brain Biopsy | ND | No | Yes, Immunomodulation therapy | No |
| 28* | 11-15 | F | Healthy | No | Sz, progressive HP | Left mesial temporal sclerosis | EBV (blood) | Brain Biopsy | ND | No | Yes, Immunomodulation for atypical Rasmussen's | No |
| 29* | 1-5 | F | Status post HSV encephalitis | No | Sz, lethargy, increased ICP | Confluent new signal change with leptomeningeal enhancement in both frontal lobes, the peri-rolandic regions and thalami. Residual encephalomalacia and hemosiderin staining | ND | Brain Biopsy | HSV | Yes (chronic HSV antiviral suppression) | Yes, Immunomodulation for post-infectious granulomatous condition + HSV antivirals | No |
| 30* | 6-10 | M | DID | Yes | Sz, enc | Multifocal cortical-subcortical ring enhancing lesions | EBV (blood) | Brain Biopsy | ND | No | Yes, Targeted therapy for atypical PRES | Yes |
| 31* | 1-5 | F | Healthy | No | Ataxia. DA, HP, hypertonia, speech regression, bulbar abn | Right cerebral atrophy | ND | Brain Biopsy | ND | No | Yes, Immunomodulation therapy for undiagnosed neuroinflammatory condition | No |
| 32* | 11-15 | F | Healthy | No | Ataxia, myoclonus, DR, oromotor apraxia | No abnormalities | ND | Brain Biopsy | ND | No | No | No |
| 33* | 6-10 | F | Healthy | No | Enc, HP | Hydrocephalus with diffuse leptomeningeal enhancement | ND | Brain Biopsy | M. tuberculosis | Yes, (targeted TB treatment) | Yes, Targeted anti-infective treatment | No |
| 34 | 11-15 | F | Healthy | No | Sz, enc | No abnormalities | ND | CSF | ND | No | Yes, Immunomodulation therapy for FIRES | No |
| 35 | 11-15 | F | IID, status post HSCT (10 years prior) | No** | Gait disturbance, hand tremor | No abnormalities | ND | CSF | ND | No | No | No |
| 36 | 1-5 | M | TD, status post thymus transplant | Yes | Sz | No abnormalities | SARS-CoV-2 (NPA) | CSF | ND | No | Yes, Immunosuppression for post-infectious immune-dysregulation | No |
| 37 | 6-10 | F | HM | Yes | HP | Multi-territorial acute infarcts, with nodular leptomeningeal enhancement | ND | CSF | ND | No | No | Yes |
| 38 | 11-15 | F | Vein of Galen malformation, status post embolisation | No | Enc, HA | Focal diffusion restriction along the globus pallidus with subarachnoid haemorrhage | ND | CSF | ND | No | No | No |
| 39 | 1-5 | M | SCID, status post HSCT | Yes | Myoclonus, enc | Meningoencephalitis with ventriculitis | CMV (blood) | CSF | CMV | Yes, (foscarnet, ganciclovir, letermovir) | Yes, Diagnostic confirmation and targeted CMV treatment | No |
| 40 | 6-10 | F | HM | Yes | HA | Right pontine diffusion restriction and left hippocampal swelling | ND | CSF | ND | No | Yes, Chemotherapy for CNS malignant spread after CNS infection ruled out | No |
| 41 | ≥16 | M | DID, status post HSCT (15 years prior) | Yes† | Descending paralysis, enc, myoclonus | Extensive cortical signal change with diffusion restriction in the left frontal, insular and parietal regions with further involvement of the left lentiform nucleus and the right peri-rolandic cortex and cerebellum | ND | Brain Biopsy | TTV, Avian orthoavulavirus | Yes (ribavirin, nitazoxanide, favipiravir) | Yes, Directed antivirals with concurrent immune-suppression of CNS HLH | No |
| 42 | 1-5 | M | SCID, status post HSCT | Yes | Gait disturbance, ptosis, weakness | No abnormalities | Rhino/enterovirus (NPA), EBV (blood) | CSF | TTV | No | Yes, Targeted Myasthenia Gravis Treatment | No |
| 43 | <1 | F | BMF syndrome | Yes | Abn movements, neck stiffness | Progressive atrophy and subdural effusions | SARS-CoV-2 (NPA) | CSF | ND | No | No | No |
| 44 | 11-15 | M | IBD, large vessel vasculitis | Yes | Sz, enc | Extensive white matter signal changes throughout with microhaemorrhages and nodular enhancement | CMV (blood), SARS-CoV-2 (NPA) | Brain Biopsy, CSF | ND | Yes | Yes, Change in Immunosuppression regimen | No |
| 45 | 1-5 | M | SCID, status post HSCT | Yes | Sz, DR, speech delay, hearing/vision loss | Multifocal areas of enhancing signal abn in the grey and white matter involving the cerebral hemispheres bilaterally, particularly the basal ganglia and parietooccipital lobes | ND | Brain Biopsy | Mumps virus (vaccine strain) | No | Yes, IVIG & reduction of immunosuppression | Yes |
| 46* | <1 | F | TD | Yes | Sz | Surface microhaemorrhages, cerebral underdevelopment | Rotavirus, norovirus (stool), BCGosis, Rhino/enterovirus (NPA) | CSF | ND | No | No | Yes |
| 47 | 1-5 | F | Healthy | No | Sz, increased ICP | Extensive craniospinal leptomeningeal enhancement, left MCA ischaemic infarcts | SARS-CoV-2 (NPA) | Spinal Meningeal Tissue | ND | Yes (targetted TB treatment) | Yes, Diagnostic confirmation and targetted anti-infective treatment | No |
| 48 | 1-5 | M | CID, status post HSCT (X2) | Yes | Increased ICP | No abnormalities | ND | CSF | HHV6(B) | No | No | No |
| 49 | 1-5 | M | HM | Yes | HP, enc, sz | Multiple ring enhancing lesions in left frontal and parietal lobes | aspergillus fumigatus (CSF 18S) | Brain biopsy | Aspergillus fumigatus | Yes (antifungal) | Yes, Exclusion of alternative/concurrent diagnosis (ie. toxoplasma) | No |
| 50 | 6-10 | M | sJIA | Yes | Sz | White matter changes in centrum semiovale | norovirus (stool), adenovirus (stool), entero/rhino (NPA) | CSF | ND | No | No | No |
| 51 | ≥16 | M | IBD, status post HSCT | Yes | limb weakness/altered sensation, areflexia | Global atrophy | ND | CSF | ND | No | NA, Results post-mortem | Yes |
| 52 | 1-5 | F | SCID, status post HSCT | Yes | Sz | Global atrophy | rhinovirus (NPA) | CSF | ND | No | No | No |
| 53 | 6-10 | F | CID | Yes | HP | No abnormalities | ND | CSF | ND | No | No | No |
| 54 | 1-5 | F | Undefined autoinflammatory disorder | Yes | Sz | Limbic enc | ND | CSF & Brain Biopsy | ND (CSF), ND (Brain Biopsy) | No | Yes, Change in immunomodulation therapy | No |
| 55 | 1-5 | M | SCID, status post HSCT | Yes | Enc | Global atrophy | BCGosis, PJP pneumonia | CSF | ND | No | No | No |
| 56 | 1-5 | M | IID | Yes | Sz, aphasia | Diffuse cortical/subcortical enhancing signal abn in temporal lobes | bacillus spp (blood) | CSF | ND | No | Yes, Change in immunomodulation therapy | No |
| 57 | 6-10 | F | Healthy | No | Enc, HP, Sz, hallucinations | Multifocal infarcts in right MCA and PCA territories | HHV6 (blood, CSF, saliva) | Brain biopsy | HHV6(B) | No | Yes, Treated for alternative diagnosis in the absence of infection (PRES) | No |
| 58 | 6-10 | F | Healthy | No | HP, ataxia, CN, DA | Global atrophy, focal pontine lesion | ND | Brain Biopsy | ND | No | Yes, Treatment of alternative diagnosis (CNS malignancy) | Yes |
| 59 | 11-15 | M | DID | Yes | Limb weakness, hyporeflexia, sz | Extensive confluent white matter and thalamic signal abn with punctate enhancement | HHV6(B) (Blood, CSF), astrovirus (stool), rhino/enterovirus (NPA), parvovirus (brain biopsy), Ecoli, klebsiella, enterococcus (dural matter) | CSF & Brain Biopsy | HHV6(B) & TTV (CSF), HHV6(B) (Brain biopsy) | Yes (foscarnet, IVIG) | Yes, Change in goals of care, salvage antiviral therapy | Yes |
| 60 | 1-5 | F | Healthy | No | Sz | Cerebellar hemispheric swelling with microhaemorrhages, and further restricted foci in the cerebral hemispheres | HHV6(B) (blood, brain) | Brain biopsy | ND | Yes (ganciclovir) | Yes, Change in goals of care, treatment of alternative diagnosis (CNS malignancy), salvage antiviral therapy | No |

### Patients CMg results

Most patients had one sample sent for metagenomics, while five (Cases 6, 10, 44, 54, 59) had both CSF and brain biopsy CMg (**Table 2**). Two patients had CMg analysis on both neurological and non-neurological tissue samples (lung, liver) both of which were non-contributory to a diagnosis. One patient had CMg sequencing of spinal meningeal tissue (Case 47). The fastest time to result is seven days, however due to constraints with batch processing the target TAT is 2 weeks. There is variability around the target TAT due to staffing and equipment constraints and confirmatory testing where required; automation of the laboratory workflow and an increase in clinical demand is likely to facilitate additional staffing and equipment resources, therefore improving the TAT. RNA degradation was highest in the one FFPE brain biopsy (RIN score=1), while for the CSF samples, RNA was out of range and always processed. 1/5 brain samples with degraded RNA, defined as RIN score of <5, were positive for a pathogen (case 49, RIN score=4.7), compared to 7/24 brain biopsies with intact or partially degraded RNA. 12 cases with brain biopsies did not have RIN score information recorded, with four of these positive for a pathogen. Rates of pathogen detection were highest for the brain biopsy material (29%) compared to CSF (15%), however the difference was not statistically significant (**Figure 3A**, Fisher’s exact test p=0.24). Microbes of unknown clinical significance were more frequent in CSF (12%) compared to brain (3%), **Figure 3A**). Additionally, a greater diversity of pathogens was identified in the brain biopsies (**Table 3**).

**Table 3:** (a) Pathogens detected in 14 patients and respective CMg read counts (b) Microorganisms of unknown clinical significance detected in 7 patients and respective CMg read counts*

| Sample | Pathogens detected | Number of cases | Pathogen read counts |
| --- | --- | --- | --- |
| **Brain** | Astrovirus VA1/HMO-C | 2 | 46 (case 1) & 4,900,000 (case 10) |
|  | Coronavirus OC43 | 1 | 997,453 (case 4) |
|  | Mumps virus (vaccine strain) | 1 | 77,624 (case 45) |
|  | Aspergillus fumigatus | 2 | 34 (Case 9) & 3135 (case 49) |
|  | Mycobacterium Tuberculosis | 1 | 26 (case 33) |
|  | Avian orthoavulavirus | 1 | 1,113,367 (case 41) |
|  | HHV6B | 1 | 25,122 (case 59) |
|  | EBV | 1 | 38,200 (case 3) |
|  | HSV | 1 | 2 (case 29) |
| **CSF** | Adenovirus | 2 | 4,300,000 (case 11) & 14 (case 14) |
|  | CMV | 1 | 25,766 (case 39) |
|  | HHV6B | 1 | 10 (case 59) |

b.
| Sample | Microorganisms of unknown clinical significance | Number of cases | Microorganisms read counts |
| --- | --- | --- | --- |
| **Brain** | HHV6B | 1 | 72 (case 57) |
|  | TTV | 1 | 165 (case 41) |
| **CSF** | HHV6B | 1 | 133 (case 48)) |
|  | HHV-7 | 1 | 6 (case 7) |
|  | TTV | 3 | 2,254 (case 11) & 50 (case 42) & 213 (case 49) |

\* TTV was usually identified in cases with pathogens detected in the same clinical sample (3/4)
CMg: clinical metagenomics; CMV: cytomegalovirus; EBV: Epstein-Barr virus; HHV: human herpes virus; HSV: herpes simplex virus; TTV: torque teno virus

**Figure 3.**
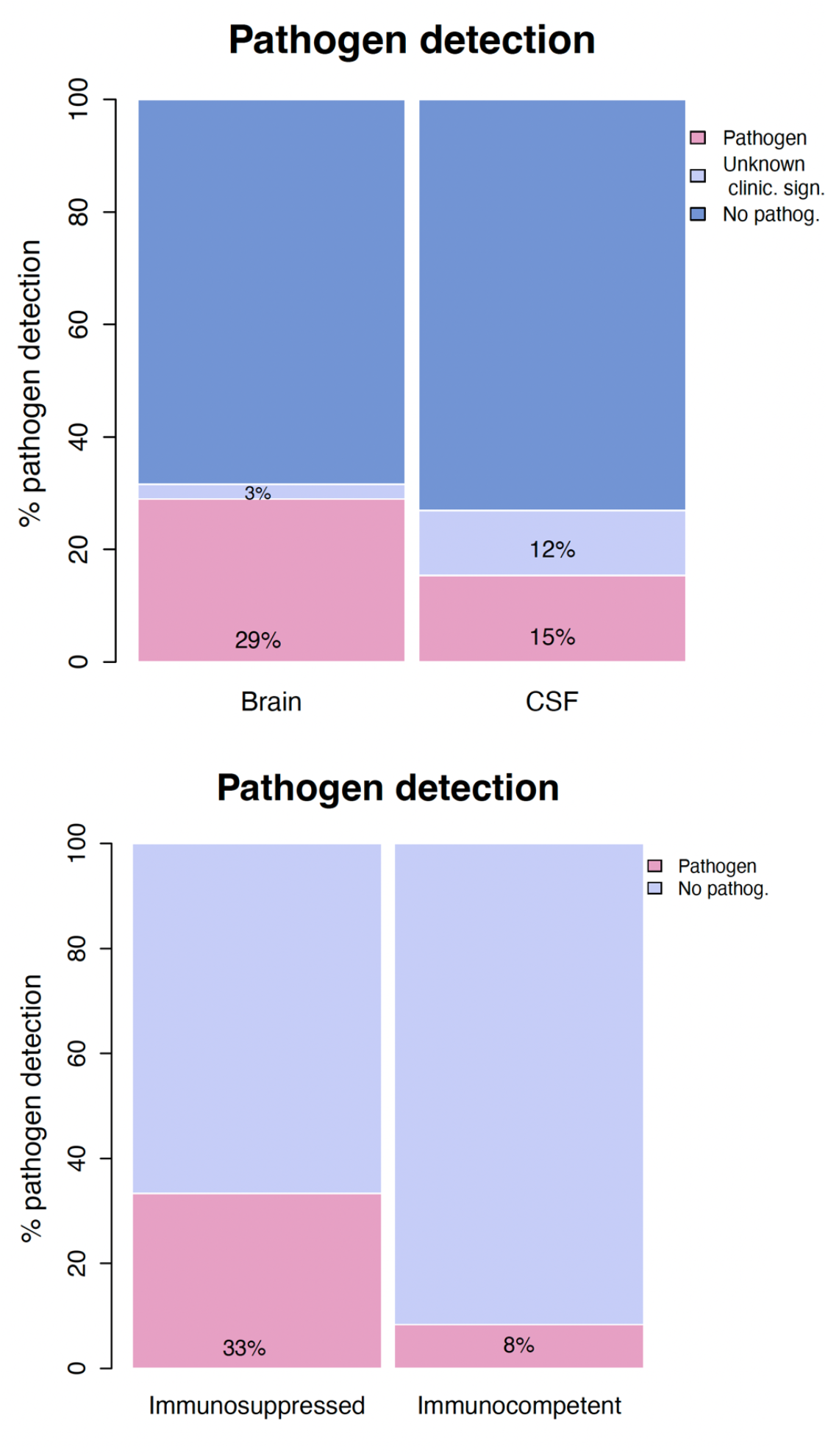
Pathogen detection rate per A. clinical specimen and B. patients’ immune system status

In the 60 patients described here, we reported a positive result in 18 (30%), including 14 patients (23%) where the causative pathogen was identified by CMg (**Tables 2 & 3**). In two patients we detected microorganisms of unknown clinical significance in the CSF (Human Herpes Virus (HHV)-7 and Torque Teno Virus (TTV), Cases 7 and 42 respectively, **Table 3**). In three patients (Cases 48, 57, 59) we detected HHV-6B, however in only one (Case 59) it was deemed to be of clinical importance. Where a novel or unexpected pathogen was detected, or when the significance of a detected pathogen was uncertain, we undertook additional confirmatory testing when possible, including PCR, serology and Immunohistochemistry (IHC) of brain tissue (cases 1, 4, 10, 41 and 45)^9,10,12^.

Two patients had pathogens identified in their brain tissue that had not previously been known to cause encephalitis: Coronavirus OC43 (Case 4)^9^ and Mumps vaccine strain (Case 45)^10^, while Avian Orthoavulavirus (Case 41) and Astrovirus VA1/HMO-C (Case 1)^12^ had been detected in cases of encephalitis only once before^13,14^. Cases 10 (Astrovirus VA1/HMO-C), 41 (Avian Orthoavulavirus), 45 (mumps vaccine strain^10^) and 29 (HSV-1) although confirmed as positive by PCR of brain tissue, were PCR negative for their respective pathogen in all other specimens tested, including CSF, blood, respiratory secretions, and stool. Case 29 had been diagnosed with and treated for HSV-1 encephalitis 10 months earlier (CSF PCR positive). With symptomatic re-presentation, brain biopsy revealed three HSV-1 reads in the RNA-seq (no DNA-seq was available at the time) which was confirmed by HSV-1 PCR of brain tissue (Ct=30).

For 8/14 patients in whom viruses were detected (Cases 3, 4, 10, 11, 39, 41, 45, 59) including 6/10 (60%) with a positive brain biopsy and 2/4 (40%) with positive CSF, full viral genome sequences were obtained (**Table 3**). This was not achieved for the remaining three virus-positive cases; neither for the three cases in whom larger bacterial and fungal genomes had been detected.

Overall, of the 36 immunocompromised patients, 12 (33%) were positive for a pathogen, in contrast to 2/24 (8%) patients classified as having no previous immunocompromising medical condition (**Figure 3B**, Fisher’s exact test p=0.03).

### MDT change in management

Of the 14 patients where the pathogen was identified by CMg, seven had a previously undetected pathogen, (Cases 1, 4, 10, 33, 41, 45, 48, **Table 2**). In the remaining seven patients (Cases 3, 9, 11, 14, 39, 49, 59 (**Table 2**) CMg confirmed a pathogen previously identified elsewhere as the cause of their encephalitis. Results for three cases (Cases 1, 4, 51) were only available post- or peri-mortem precluding clinical intervention (**Table 2**).

Based on CMg results, the MDT recommended changes to clinical management in 74% (n=42) of the 57 cases for whom results were obtained antemortem. This included 24/33 (73%) of the immunocompromised patients and 18/24 (75%) of the immunocompetent referrals. In 13 cases (22%), the CMg results together with MDT input led to initiation of targeted anti-infective therapy with reduction of broad-spectrum antimicrobials (**Table 2**). In three patients (cases 10,11, 41) the MDT sanctioned the use of experimental repurposed antiviral agents; all three cases are alive in follow up (Breuer personal coms) (**Table 2**). Salvage antifungal therapy was recommended by the MDT in four patients (cases 9, 16, 19, 49), 3 based on CMg/PCR findings and one based on neuroimaging findings alone.

In four cases (Cases 10, 33, 41, 45), where prior to CMg, the diagnostic balance had leant towards an inflammatory pathogenesis, the discovery of a pathogen led to reduction in immunosuppressive and anti-inflammatory agents. In one immunocompetent patient (case 29) the MDT recommended concurrent immunomodulation (steroids) with chronic HSV-1 antiviral suppression. Conversely, in 21 (35%) cases, the absence of a pathogen detected by CMg led the MDT to recommend starting, increasing, or changing immunomodulation/immune-mediated therapies and in one patient (case 18) the absence of infection detected by CMg informed MDT recommendation to proceed to HSCT. In eight (13%) cases, in whom CMg did not find a pathogen, were continued on targeted or symptomatic therapy for non-infectious or inflammatory pathology (atypical presentations of primary neurological conditions eg. posterior reversible encephalopathy syndrome [PRES], myasthenia gravis, or malignancy) with increased assurance against an infective aetiology (cases 7,12,17,30,40,42,57,58). For five patients (cases 10,14,19,59,60), the CMg result led to a change in goals of care, with greater emphasis on palliation, although initially salvage anti-infective therapy was considered in four.

Overall, 45/57 patients with an antemortem CMg result are alive in follow-up, including eight in whom new specific antimicrobial treatment was started following pathogen detection. Nonetheless, mortality was high, with 15/60 patients (12/57 with an antemortem diagnosis) dying, including eight despite neuro-infection MDT informed modification to management. 12/15 patients who died were immunocompromised at the time of referral with a median age of five years. Two of the three immunocompetent patients were later found to have a malignancy affecting the CNS. Five immunocompromised patients (cases 1, 4, 14, 45, 59) had a clinically relevant pathogen detected and barring case 59, it was concluded that the infective encephalitis had been the main contributor to their death. For three of these patients, the CMg pathogen detection was peri-mortem or post-mortem. The remaining seven cases in whom a pathogen was not detected, died from complications of their underlying disease.

## Discussion

Identifying the aetiology of meningoencephalitis in complex patients referred to tertiary or quaternary centres is frequently urgent and can be very challenging. Neurological (**Table 2**) and neuroradiological features (**Supplementary Figure 1**) are diverse and as we show here, frequently cannot distinguish between infectious or non-infectious aetiologies. We have shown in this large retrospective observational case-series, that an integrated CMg and clinical neuro-infection MDT can significantly improve diagnostic yield when investigating medically complex patients. This in turn can have real time influence on potentially lifesaving management decisions. This is particularly true for immunocompromised hosts who are at higher risk of unusual CNS pathogens, which may not be detected by standard investigations. Immunocompromised patients are also at risk of CNS immune dysregulation, with, or as a sequela of active infection, and who are known to suffer worse outcomes when untreated^15,16^.

The most evident benefit of CMg is the identification of pathogens that could not otherwise be detected even by the extended diagnostic tests available. In our cohort, two patients had pathogens in their brain tissue that had not previously been known to cause encephalitis: coronavirus OC43 (Case 4)^9^ and mumps vaccine strain (Case 45)^10^, while cases of avian orthoavulavirus (Case 41), and astrovirus VA1/HMO-C (Case 1)^12^ had only been detected once before^13,14^ and therefore targeted diagnostic tests were not available or considered relevant. For cases 41 and 45, negative CSF results had initially led to escalation of immunosuppressive and immunomodulatory treatments for presumed inflammatory aetiology, with the CMg findings resulting in modification to planned therapy. Neurotropic astroviruses have since been reported in several cases of undiagnosed encephalitis occurring in immunocompromised individuals^17–20^ while coronavirus OC43 has been identified in two more cases^21,22^. Both should arguably now be included as part of routine testing in immunocompromised individuals with encephalitis of unexplained aetiology, even if CMg is not undertaken.

Four cases (Cases 41, 45, 10, 29) were positive for the causative agent only in brain tissue, but not in any other specimens tested, including CSF. The discovery of an unexpected and/or novel pathogen, which otherwise would likely remain undiagnosed and fatal, may lead to anti-infective directed therapies. While treatment options, particularly for viral causes of encephalitis remain few, the potential use of repurposed antiviral agents with broad spectrum anti-RNA viral activity is being actively investigated^23,24^. Personalised, experimental antiviral treatment has already been used successfully in patients with encephalitis^23^ including one patient with neurotropic astrovirus (Breuer personal communication).

Where known pathogens were found, CMg results still informed treatment decisions. In case 9, who had documented infection with aspergillus in non-CNS tissue culture and group B streptococcus in blood culture prior to developing encephalitis, CMg found only aspergillus in brain tissues, with no other pathogens detected, allowing targeted antifungal treatment. In Case 12, CMg detected adenovirus in brain coinciding with the only pathogen found by PCR in blood, providing support for targeted antiviral treatment with brincidofovir alone. Furthermore, while EBV and CMV viraemias were frequent in our patient cohort, with five and four cases respectively, a positive CMg finding in the brain biopsy (case 3) or the CSF (case 39) resulted in recommendation for targeted EBV PTLD treatment and targeted CMV treatment respectively. In contrast, a negative CMg result, particularly since 2019, with both DNA-seq and RNA-seq undertaken to achieve universal pathogen detection, provided increased confidence in a likely inflammatory aetiology, primary or post-infectious, and resulted in MDT recommendations to start or escalate immunomodulatory and/or immunosuppressive therapies.

Change in management recommended by the MDT could be broadly grouped into four categories (1) pathogen directed therapy (2) immunomodulation (3) targeted therapy for non-infective/non-inflammatory pathology 4) re-evaluation of goals of care. CMg results received antemortem, led to one or more of the above decisions being applied to at least 73% (24/33) of immunocompromised patients. However, not all MDT-recommended treatment changes were due to CMg results alone, and co-ordinated multidisciplinary specialist input was of primary importance. Close communication between laboratory, medical and surgical teams throughout the process is essential to optimise diagnostic yield and in interpretation of clinical implications of both positive and negative CMg findings. In depth knowledge of test sensitivity and specificity, and biological plausibility of detected pathogens contributing to presentation is key. Increasing use of CMg, together with evolution of the MDT with time, has led to institutional learning and changes in the patient clinical pathway. CMg, initially of CSF, is now incorporated early into the diagnostic pathway, especially for immunocompromised patients. Brain biopsy with routine storage of a portion in RNAlater to allow CMg if required is carried out where undiagnosed infection remains a possibility. This approach is supported by the excellent safety record of brain tissue biopsy in children^25^ and at GOSH in particular^26^.

There are several caveats to this report. First, the clinical interpretation of CMg is highly dependent on the test performance characteristics. While the CMg service at GOSH has been thoroughly and repeatedly validated for use, it has evolved over the years with improvements in laboratory and bioinformatic pipelines introduced as wet lab and bioinformatics advancements became available.

Contextualisation of CMg results was required, usually when microorganisms are found at low levels or do not fit with the clinical presentation. Two examples are TTV and HHV-7, found by CMg in our cohort (Case 7, 42) and interpreted to be of unknown clinical significance. TTV is a common commensal agent, its seroprevalence varying according to geographical location and is estimated globally to be present in blood from 50% of the general population^27^. In our cohort, we have infrequently detected it in CSF and once in brain tissue, and its clinical significance is unknown^28^. HHV-7 persists in most adults^29^, has not been associated with disease and may reactivate in immunocompromised individuals^29^. Similarly, where HHV6 is detected, it is important to exclude chromosomal integration of virus and/or sample contamination with viraemic blood (cases 48, 57) from cases where the virus is likely to be the primary or a contributing infection (case 59). Finally, it is important to recognise the limits of CMg. Our current methods, although validated as equivalent to targeted real-time PCR for CSF, are only as sensitive as 16S rRNA gene PCR and less sensitive than targeted real-time PCRs for detection of DNA pathogens in tissue. For this reason, we have a lower threshold for confirmatory PCR of brain tissue where any reads from a DNA pathogen are detected by CMg (e.g Case 29).

While broad based, unbiased testing can be expensive and is currently limited to centres with laboratory capacity and expertise in metagenomics practices, it does overcome the constraints that small pediatric clinical specimen volumes place on testing for multiple pathogens, particularly where a pathogen is rare or unexpected and is therefore unlikely to be part of a routine PCR testing algorithm. Increased funding for and the development of referral centres of excellence in CMg that can process larger numbers of samples with quicker turnaround time are likely to decrease cost and foster further translation of specialised CMg knowledge. Institutional surgical expertise for rapid invasive CNS sampling is also required; the GOSH MDT practice is now to recommend earlier consideration of brain biopsy. Furthermore, the multidisciplinary nature of such a team of specialists requires commitment in terms of time allotment and necessitates considerable organisation to formulate comprehensive MDT pathways and procedures. Likewise, the MDT must be responsive in terms of acuity, as complex, undefined meningoencephalitis patients require prompt intervention without administrative delay.

Our analysis is limited by the retrospective single centre, observational nature of the study with limited patients and heterogenous presentations and lack of external validation or biologic controls from neuropathologic samples without meningoencephalitis not seen through the MDT. MDT outcomes can be subjective, although we attempted to overcome this with two independent clinicians analysing patient data. Similarly, MDT workflow patterns and CMg analysis changed over time, limiting standardisation of data collection. Shifting expertise within the MDT over time, as knowledge increased and with diagnostic advancements, is also likely to have altered MDT decision making. We cannot fully guarantee that all MDT recommendations were followed by the primary treating physicians as outlined in the MDT outcomes although we did make every attempt to verify through chart reviews, personal communications, and follow-up evaluations through the existing neuro-infection MDT pathway.

In summary, we report a combined clinical neuro-infection MDT service that has successfully integrated CMg into the diagnostic pathway for complex undiagnosed CNS presentations where infection remains in the different although unconfirmed despite extensive diagnostics. Based on the findings described here, the clinical practice for all patients with undiagnosed meningoencephalitis at GOSH, but particularly those who are immunocompromised, now includes early discussion at the MDT, and routine recommendation of CSF CMg. For patients with negative microbiological results with infection still in the differential diagnosis, CMg of brain tissue is undertaken where possible, after careful consideration of risk/benefit. Importantly, the combined MDT and CMg service are now offered to clinical groups across the world, with over half of specimens for CMg now referred from outside GOSH. Our data demonstrate that both positive and negative CMg results can be of great utility in clinical decision making particularly in immunocompromised individuals. Improved turnaround times and decreased cost of CMg, especially in the context of high healthcare costs for undiagnosed cases of encephalitis and the development of new personalised treatments, should encourage routine adoption into clinical practice. Patient selection remains fundamental and diagnostic stewardship of results by an MDT essential.

## Data Availability

All data produced in the present study are available upon reasonable request to the authors

## Ethics

Metagenomic analysis were carried out by the routine diagnostic service at Great Ormond Street Hospital. Additional PCRs, Immunohistochemistry on samples received for metagenomics (not discussed in the paper in detail) are part of the Great Ormond Street Hospital (GOSH) protocol for confirmation of new and unexpected pathogens. The use for research of anonymised laboratory request data, diagnostic results and residual material from any specimen received in the GOSH diagnostic laboratory was approved by UCL Partners Pathogen Biobank under ethical approval granted by the NRES Committee London-Fulham (REC reference: 17/LO/1530).

Demographic and clinical information on cases included in this study were obtained according to the clinical audit with registration number: 3583.

The case IDs in the paper are anonymised IDs that cannot reveal the identity of the study subjects and are not known to anyone outside the research group, such as the patients or the hospital staff.

## Acknowledgements

We would like to recognise the large group of junior doctors, clinical nurse specialists, laboratory scientists, and specialist physicians that contribute their expertise to the GOSH neuro-infection MDT. We would like to specifically thank Drs Louis Grandjean, Seilesh Kadambari, Nigel Klein, Ashwin Pandey, James Soothill, Claire Booth, Winnie Ip, Catherine O’Sullivan, Reem Elfeky, Waseem Qasim, John Hartley, Cheryl Hemingway, and Yael Hacohen for their contributions. In addition, we would like to acknowledge the patients and their families for their valued contributions to the clinical MDT and the advancement of translational diagnostics.

S.M is funded by a W.T. Henry Wellcome fellowship (206478/Z/17/Z). J.B receives funding from the National Institute of Health Research UCL/UCLH Biomedical Research Centre. TJ is grateful for funding from the Brain Tumour Charity, Children with Cancer UK, Great Ormond Street Hospital Children’s Charity, Olivia Hodson Cancer Fund, Cancer Research UK and the National Institute of Health Research.

All research at Great Ormond Street Hospital NHS Foundation Trust and UCL Great Ormond Street Institute of Child Health is made possible by the NIHR Great Ormond Street Hospital Biomedical Research Centre.

The views expressed are those of the author(s) and not necessarily those of the NHS, the NIHR or the Department of Health.

## Contributors

JP, SM, JB, AB, MK conceived and designed the study, JP, JB & SM wrote the manuscript, JP, NR, DC, GD collated the patients’ clinical data, JP, JHassell assessed and categorised the MDT outcomes, JRB developed the wet-lab CMg methods, JRB, LA, AL, JCDL, AK, DS processed samples and generated the CMg data, SM developed the computational methods, SM, CV, NS analysed the CMg data, SM, NS, JRB, LA, SD, JHatcher, KH and JB interpreted the CMg data, MK, AB, DS, KMoshal developed the initial framework for the neuro-infection MDT, MK and JHassell are the MDT neurology leads, AB is the infectious diseases MDT lead, JHatcher the microbiology MDT lead, MAK and AJJW the clinical immunology MDT leads, AM and TJ the histopathology/neuropathology MDT leads, JB the clinical virology MDT lead, KA the neurosurgery MDT lead, and GL the BMT MDT lead. KMankad is the neuroradiology MDT lead, interpreted radiological findings and collated radiological figures.

