## Supplementary Material for "Translating metagenomics into clinical practice of complex paediatric neurological presentations"

**Supplementary Methods**

**Untargeted metagenomics**

**Nucleid Acid Extraction**

Up to 20 mg of biopsy is lysed with 1.4mm ceramic, 0.1mm silica and 4mm glass beads, prior to DNA and RNA purification using the Qiagen AllPrep DNA/RNA Mini kit as per manufacturers’ instructions, with a 30 µl elution volume for RNA and 50 µl for DNA. Biopsies that have not been collected into RNA*Later* are infused with RNA*Later*-ICE prior to nucleic acid extraction. Formalin fixed paraffin embedded (FFPE) biopsied are deparaffinized with Qiagen deparaffinization solution, lysed with 0.1 mm and 1 mm glass beads and extracted using the Qiagen EZ1 RNA tissue kit and DNA blood kit with 50 µl elution volumes.

500 µl of CSF samples are lysed with 0.1 mm and 1 mm glass beads prior to DNA and RNA purification using the Qiagen EZ1 virus mini kit, without carrier RNA, and a 60 µl elution volume.

**Metagenomics library preparation and sequencing**

RNA from CSF with RNA yield >2.5 ng/µl and from biopsies underwent ribosomal RNA depletion and library prep with KAPA RNA HyperPrep kit with RiboErase, according to manufacturer’s instructions. RNA from CSF with RNA yield <2.5 ng/µl did not undergo rRNA depletion prior to library prep. DNA from CSF with DNA yield >1 ng/µl and from biopsies underwent depletion of CpG-methylated DNA using the NEBNext® Microbiome DNA enrichment kit, followed by library preparation with NEBNext Ultra II FS DNA Library Prep Kit for Illumina, according to manufacturer’s instructions. DNA from CSF with DNA yield <1 ng/µl did not undergo depletion of CpG-methylated DNA prior to library prep. Sequencing was performed with a NextSeq High output 150 cycle kit with a maximum of 12 libraries pooled per run, including controls.

**Negative/Positive controls**

Every batch of samples is accompanied by a control sample containing feline calicivirus RNA and cowpox DNA which was processed alongside clinical specimens, from nucleic acid purification through to sequencing. All specimens and controls are spiked with MS2 phage RNA internal control prior to nucleic acid purification.

**Limit of detection and validation**

To establish the limit of detection for DNA and RNA microorganisms in brain tissue and CSF, we spiked mock tissue with dilution series of cowpox DNA (CMg DNA-seq detected to Ct=31). We also spiked mock tissue with dilution series of feline calcivirus RNA (CMg RNA-seq detected to Ct=39). Known positive samples detected by metagenomics can be seen in **Supplementary Table 2**.

**Bioinformatics**

**Preprocessing pipeline**

Briefly it consists of trimming adapters and low-quality ends (TrimGalore ^30^-version 0.3.7), filtering human sequences (Bowtie2^31^, version 2.4.1 29, GRCH38 p.9 and megaBLAST, version 2.9.0), removing low quality and low complexity sequences (PrinSeq^32^, version 0.20.330). For RNA-seq, ribosomal RNA sequences are removed using a similar 2 step-approach (Bowtie2 and megaBLAST). Prior to taxonomic classification, we perform nucleotide and protein similarity searches (megaBLAST and DIAMOND^33^,version 0.9.30) against custom reference databases (RefSeq nucleotide and protein collections of viruses, bacteria, fungi, parasites and human, downloaded March 2020).

**Supplementary Tables**

*Supplementary Table 1 - Comprehensive Neuro-infection investigations routinely considered and recommended by MDT*

| Infection Investigations | Neurology & Other Investigations |
| --- | --- |
| Bacteria   - Blood culture - Urine culture - CSF bacterial culture - CSF specific PCR: e.g. N*eisseria meningitidis, Streptococcus pneumoniae, Streptococcus agalactiae, Enterobacteriaceae spp* - CSF and blood: 16S PCR - Syphilis serology - Other bacterial serology/PCR as clinically indicated based on epidemiology/exposure risk (eg. Lyme serum:intrathecal index and/or PCR) | CSF   - CSF cell count, protein, glucose - CSF oligoclonal bands (paired with serum)   With MDT Consideration, CSF:   - Neopterins - Cytokines - Immunophenotyping - NMDA encephalitis antibodies - Paraneoplastic antibodies - Cytospin   With MDT consideration, blood:   - Antineuronal antibodies (eg. MOG, NMDA, Aquaporin 4)   With MDT consideration, histology:   - Brain Biopsy (after MDT risk/benefit analysis) |
| Viral   - Blood specific PCR: e.g. HSV, VZV, enterovirus, parechovirus, adenovirus, EBV, CMV, parvovirus, HHV6/7 - CSF specific PCR: HSV, VSV, enterovirus, parechovirus, , HHV 6/7, EBV, CMV, adeno, astrovirus - HIV serology/PCR - Nasopharyngeal multiplex respiratory PCR - Stool: enterovirus, parechovirus, multiplex gastroenteritis virus PCR panel - Other viral serology/PCR as clinically indicated based on epidemiology/exposure risk eg. measles IgG (serum:intrathecal index) | Radiology   - MR Brain +/- spinal imaging (additional MR sequences including contrast, time of flight angiography, spectroscopy, angiography as guided by neuroradiology) - Chest, abdominal, pelvic imaging as clinically indicated   With MDT Consideration:   - PET scan |
| Fungal   - Blood fungal culture - CSF fungal culture - Blood and CSF: ITS PCR - Fungal markers: B-D glucan, galactomannan - Endemic fungal serology/urinary antigens as indicated | Electrophysiology   - EEG - Nerve conduction studies as clinically indicated - EMG studies as clinically indicated |
| Mycobacterial   - Interferon gamma release assay - Mycobacterial cultures and PCR (blood, respiratory, bone marrow) - CSF: mycobacterial PCR (pan-mycobacterial) and culture | Immunology & Metabolic   - Quantitative & Functional immunologic testing for inborn errors of immunity as clinically indicated - Investigations for inborn errors of metabolism as clinically indicated - Toxicology as clinically indicated |
| Parasites   - Blood/CSF: Toxoplasma serology/PCR - Malaria (as clinically indicated) - Other parasitic testing as indicated based on epidemiology/exposure risk (serology and/or specific PCR) | Ophthalmology   - Eye exam for chorioretinitis, uveitis, optic neuritis etc. |

CSF: cerebrospinal fluid; EEG: CMV: cytomegalovirus; electroencephalogram; EBV: Epstein-Barr virus; EMG: electromyography; HHV: human herpes virus; HIV: human immunodeficiency virus; HSV: herpes simplex virus; ITS: internal transcribed spacer; MDT: multidisciplinary team; MOG: Myelin oligodendrocyte glycoprotein; MR: Magnetic resonance; NMDA: N-methyl-D-aspartate; PCR: polymerase chain reaction; PET: positron emission tomography; VZV: varicella-zoster virus

*Supplementary Table 2: Known positives detected by metagenomic as part of validation work*

**Mock Tissue** – spiked dilution series results with model organisms

| **RNA virus (feline calicivirus)** | | **DNA virus (cowpox virus)** | |
| --- | --- | --- | --- |
| **PCR Ct value** | **Detection by mNGS (No of reads)** | **PCR Ct value** | **Detection by mNGS (No of reads)** |
| 29 | Detected (3,095) | 28 | Detected (676 reads) |
| 32 | Detected (5,412) | 31 | Detected (81 reads) |
| 39 | Detected (344) | 34 | Not detected |
|  |  | 36 | Not detected |

**Mock CSF** – spiked dilution series with model organisms

| **RNA virus (feline calicivirus)** | | **DNA virus (cowpox virus)** | |
| --- | --- | --- | --- |
| **PCR Ct value** | **Detection by mNGS (No of reads)** | **PCR Ct value** | **Detection by mNGS (No of reads)** |
| 30 | Detected (436 reads) | 28 | Detected (1329 reads) |
| 33 | Detected (1,090 reads) | 31 | Detected (127 reads) |
| 37 | Detected (87 reads) | 34 | Detected (32 reads) |
| 41 | Detected (20 reads) | 37 | Detected (11 reads) |

**Residual Tissue** – known positive residual samples (n=10) results

| **Target organism** | **PCR Ct value** | **Detection by mNGS (No of reads)** |
| --- | --- | --- |
| **RNA virus** | | |
| Astrovirus VA1/HMO-C | 31 | Detected (4.9 million) |
| **DNA virus** | | |
| VZV (FFPE) | 31 | Detected (18) |
| JC virus (FFPE) | n/a* | Detected (17,170) |
| AAV-2 (ssDNA) | 17 | Detected (1420 reads) |
| AAV-2 (ssDNA) | 18 | Detected (576 reads) |
| AAV-2 (ssDNA) | 18 | Detected (1253 reads) |
| AAV-2 (ssDNA) | 19 | Detected (267 reads) |
| HHV6 | 36 | Not detected |
| HHV6** | 27 | Detected (209 reads) |
| HHV6** | 29 | Not detected |
| HHV6** | 30 | Detected but below reportable limit (6 reads) |
| HHV6** | 32 | Detected but below reportable limit (6 reads) |
| Adenovirus** | 37 | Not detected |
| Adenovirus** | 37 | Not detected |
| Adenovirus** | 38 | Not detected |
| Adenovirus** | 42 | Not detected |
| **Bacteria** | | |
| *Rothia mucilaginosa* | n/a (16S PCR positive) | Detected (1,502) |
| BCG | 35 | Detected (10 reads *M. bovis*) |

*(histopathology +ve SV40 staining), CSF JC PCR positive (brain tissue not tested by PCR)

**same specimens as the AAV2 biopsies

**Residual CSF** – known positive residual samples (n=9) results:

| **Target organism** | **PCR Ct value** | **Detection by mNGS (No of reads)** |
| --- | --- | --- |
| **RNA virus** | | |
| Enterovirus*** | 32 | Detected (1,685 reads) |
| Enterovirus (Coxsackie B5) | 32 | Detected (128 reads) |
| **DNA virus** | | |
| Adenovirus | 20 | Detected (4.3 million) |
| Parvovirus | 29 | Detected (1,665) |
| EBV | 30 | Detected (42*) |
| HSV-1 | 30 | Detected (1,257) |
| EBV** | 35 | Detected (117) |
| HHV-6 | 36 | Not detected |
| **Bacteria** | | |
| *Enterococcus faecalis* | n/a (culture positive) | Detected (22,557) |

*by RNA sequencing only (DNA sequencing not performed on this sample)

**sub-optimal sample volume (100 µl instead of 500 µl)

*** sub-optimal sample volume (280 µl instead of 500 µl)

**SUPPLEMENTARY FIGURES** *Supplementary Figure 1. Representative images in 14 children with positive metagenomics testing*

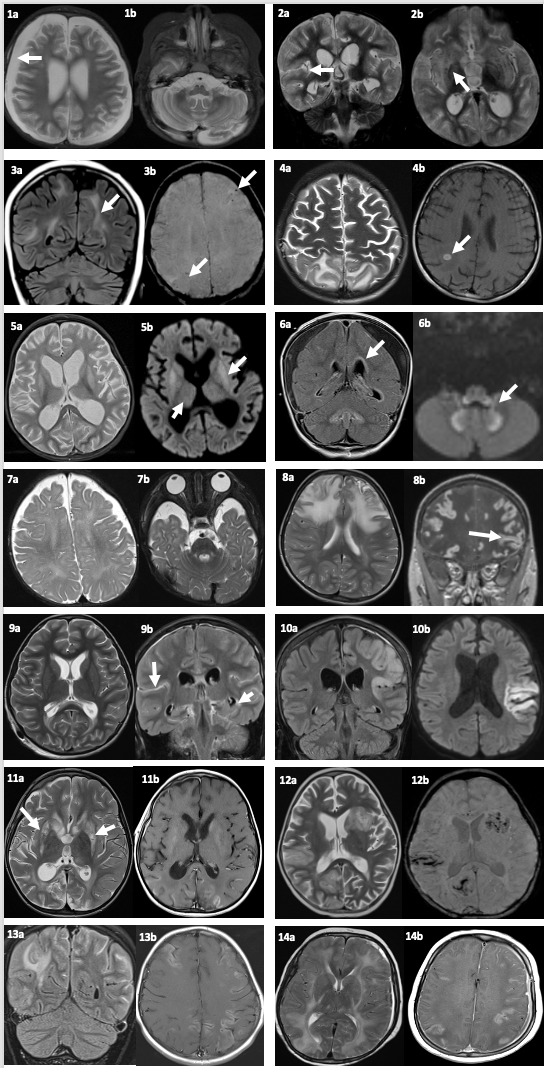

Image 1 (Case 1): progressive global atrophy with bilateral subdural effusions (arrow, 1a).

Image 2: (Case 4): progressive global atrophy, with diffuse leptomeningeal infiltration (arrow, 2a). Signal abnormality is noted in bilateral basal ganglia structures.

Image 3 (Case 7): subcortical white matter signal abnormalities and intrinsic microhaemorrhages in both cerebral hemispheres.

Image 4 (Case 9): bilateral multifocal cortical-subcortical signal changes, with micro-abscesses and micro-haemorrhages.

Image 5 (Case 10): progressive global atrophy, and bilateral symmetric signal abnormality in bilateral basal ganglia and thalami.

Image 6 (Case 11): ventriculitis, subdural effusions, and symmetric diffusion restricting changes in the brainstem nuclei, cerebellar peduncles and the dentate nuclei.

Image 7 (Case 14): global atrophy and patchy subcortical and deep white matter signal abnormality in both cerebral hemispheres.

Image 8 (Case 29): progressive bilateral asymmetric cortical-subcortical signal abnormality with cortical contrast enhancement in the backdrop of mature changes from previous herpetic infection.

Image 9 (Case 33): hydrocephalus and leptomeningeal enhancement in the brain (arrow, 9b) and spine.

Image 10 (Case 41): asymmetric cortical restriction diffusion and contrast enhancement.

Image 11 (Case 45): bilateral striatal necrosis with further contrast enhancing signal abnormality in the parietal and temporal regions.

Image 12 (Case 49): multiple bilateral lesions with patchily restricted diffusion, microhaemorrhages (12b).

Image 13 (Case 57): cortical-subcortical signal abnormality and leptomeningeal and cortical contrast enhancement.

Image 14 (Case 59): extensive confluent signal changes in the white matter of both cerebral hemispheres, and patchy contrast enhancement consistent with breakdown of the blood brain barrier.
